# Monitoring the battleground: Antimicrobial resistance, antibiofilm patterns, and virulence factors of wound bacterial isolates from patients in hospital system

**DOI:** 10.1101/2023.01.05.23284224

**Authors:** Silas O. Awuor, Richard Mariita, Eric O. Omwenga, Jonathan Musila, Stanslaus Musyoki

## Abstract

Extensive use of antibiotics in the treatment of bacterial infections has led to a challenge of antibiotic resistance, contributing to morbidity and mortality. Herein, evaluation of bacterial isolates from patients with wound discharge was performed and drug susceptibility patterns examined, with a goal of deciphering antibacterial resistance. A cross-sectional study was conducted between March to June 2022, in which samples were collected from patients with chronic wounds and were inoculated into appropriate media for identification and characterization. The bacterial pathogens were identified using standard microbiological methods. Shockingly, the majority of wound isolates showed positive growth in microbial analysis with high prevalence in male candidates. Further, *Staphylococcus aureus* 28 (20.7%) was identified as the most predominant pathogen followed by *Klebsiella spp*. 20 (14.8%), *P. aeruginosa spp*. 10 (14.8%) and lastly *E. coli* 6 (4.4%) bacteria in the wound isolates while cotrimoxazole 13 (48.1%) followed by clindamycin 7 (25.9%) and erythromycin 7 (25.9%) were the most antibacterial resistant drugs to both Gram positive and Gram-negative bacteria. Out of the four isolates, 3 (75%) isolates were able to produce the haemolysin and protease and 2 (50%) isolates were able to produce the lipase and phospholipase. The findings herein form a clinical basis for identification of antimicrobial resistance in chronic wounds that can be applied in responsible use of antibacterial in chronic wound management and as an illumination in development of more potent antibiotics for chronic wound treatment

## Introduction

The six pathogens represented in the acronym ESKAPE (*Enterococcus faecium, Staphylococcus aureus, Klebsiella pneumoniae, Acinetobacter baumannii, Pseudomonas aeruginosa*, and *Enterobacter* species) exhibit multidrug-resistance and virulence, and are attributed to a majority of nosocomial infections [1,2]. These organisms mostly infect wounds as it offers a moist, warm, nutritive environment advantageous to microbial colonization, spread, and infection [3–6]. Different bacterial species live on human skin, in the nasopharynx, gastrointestinal tract, and other parts of the body with little latent for causing disease, because of the skin being the first line of defense within the body [7, 8]. Any breach in the skin surface whether through contact with contaminated water, trauma, accident, surgical operation, or burn provides an open door for bacterial infections [7]. Consequently, wounds can be infected by a diverse array of microorganisms ranging from bacteria to fungi and parasites as well as viruses [9,10].

Unfortunately, the control of wound infections has become more challenging due to widespread bacterial resistance to antibiotics, and to a greater incidence of infections caused by methicillin-resistance *S. aureus*, polymicrobial flora [11]. Although modernization of antimicrobials in the 20^th^ and 21^st^ century was meant to transform treatment of human infections; the fast-increasing antimicrobial resistance poses a threat to mitigating infections. Antimicrobial resistance remains a dynamic risk to the management of the rising range of infections caused by microorganisms [12], hence antibacterial drugs effectiveness is reduced making the management of patients demanding and expensive inducing persistent sickness and increasing deaths on helpless patients [12]. Progress of antimicrobial resistance is a normal incidence in microorganisms which is boosted due to burden brought about by usage and mistreatment of antibiotics in organisms [13]. Lately, there has been a developing desire for identification antibiotics which are more powerful in resistant bacteria management [13]. This is because the chief common bacteria have developed resistance to common antimicrobials discovered recently [13, 14]. The absence of fresh antibiotics in the World to substitute the incompetent one conveys more urgency to the desire to guard the effectiveness of current medications, advancement and enactment of suitable methods to curb the rise and spread of antimicrobial resistance [15].

In developing countries, antimicrobial resistance poses a great trial to community health care motivating several researchers to determine the resistance profile of antibiotics against bacteria. Nevertheless, the conclusions continue to produce different opinions on the drug’s effectiveness. For instance [16, 17] in Libya like [18] in Sudan revealed sensitivity to 54% amikacin and 59% ciprofloxacin but resistance to 81% vancomycin, 75% amoxicillin, 92% streptomycin, 45% tetracycline, 26% methicillin, 65% amoxicillin, and 46% erythromycin. On the other hand, it was displayed that bacteria were highly resistant to 96% ciprofloxacin in Nigeria [19], which opposes other findings.

The increasing problem of antimicrobial resistance remains a worrying nightmare across the world prompting endless research and innovation approaches research on bacterial susceptibility and resistance to antibiotics is a top interest sometimes presenting contradicting findings. For example, in wound management, [19, 20] recognized that bacterial isolates were therapeutically responsive to amoxicillin clavulanate, meropenem, clindamycin, ceftriaxone, piperacillin-tazobactam, ciprofloxacin, vancomycin, levofloxacin, linezolid, teicoplanin, imipenem, meropenem, amikacin and levofloxacin but resistant to ampicillin, Amoxicillin clavulanate, cotrimoxazole, doxycycline and cephalosporins. Later studies [20] with similar focus presented e highly resistance to cephalosporins, amoxicillin clavulanate and imipenem with disregard to [21] findings.

## Materials and Methods

An examining study was carried out in all the samples receive at the Medical Microbiology Laboratory Department of Jaramogi Oginga Odinga Teaching and Referral Hospital (JOOTRH), Kisumu county-Kenya for the period of March to June 2022. Samples (wound pus/swab) were obtained from the patients who only attend JOOTRH both in Outpatient and Inpatient department, patient referend to the hospital were not included in this study.

Following aseptic techniques, microscopy was performed to analyze for type and morphology of bacteria. The samples were then inoculated on blood agar (BA), Mac-conkey agar (MA) and chocolate agar (CA) following previously used and established standard protocols [22]. BA and MA plates were incubated aerobically, while CA plates were incubated in anaerobic condition at 37°C for 24 to 48 hours in the incubator. All the isolates were identified by colony morphology, Gram staining reaction and the biochemical properties [22]. The antimicrobial susceptibility test of isolates was performed by the Kirby-Bauer disc diffusion method using the CLSI guidelines on Müeller-Hinton agar plates (Mast Diagnostics Ltd, Merseyside, UK) where 16 antimicrobials i.e. penicillin (25 μg), Ampicilin (10 μg), Vancomycin (15 μg), Cefoxitin (10 μg), Cefuroxime (30 μg), Oxacillin (15 μg), Ciprofloxacin (10 μg), Clindamycin (25 μg), Co-trimoxazole (30 μg), Tetracycline (15 μg), Erythromycin (10 μg), Gentamicin (30 μg), Ceftriaxone (10 μg), cefepime (30 μg), Imiperine (10 μg) and Amikacine (30 μg) will be used in this study.as outlined by Clinical and Laboratory Standards Institute (CLSI) (2022) [23]. After this isolated *S. aureus* was screened for methicillin resistance using cefoxitin disc (30 μg) as per standard guidelines provided by CLSI, (2022). MICs of ciprofloxacin and gentamicin were determined by agar dilution method [25, 26] and the guidelines of the CLSI [23]. The tests were performed by making a series of antibiotic concentrations on Mueller–Hinton agar (MHA) plates. *P. aeruginosa* spp. ATCC^®^ 12934 and *S. aureus* ATCC^®^ 29213 were used as reference strains (controls). All the data were entered in SPSS version 20 and GraphPad Prism version 9.4. Statistical analyses were done using the same software.

### Biofilm formation inhibition assay

As described by Awuor et al. [27], microtiter plate assay was performed to quantify the effect of commonly used antibiotics on the biofilm formation of *P. aeruginosa spp*. strains. The test bacteria were first inoculated on Luria-Bertani (LB) medium agar and incubated at 37 °C overnight. Then a colony was identified, picked and inoculated in 10 ml of LB broth and incubated at 37 °C overnight while shaking at 100 r.p.m. for 18 h. By use of a parafilm the flat-bottomed polystyrene tissue culture microplate was sealed for purposes of preventing medium evaporation. After 48 h incubation, the wells were carefully rinsed with double-distilled water to remove loosely attached cells. The microplate was air-dried for 1 h before adding 200 μL per well of 0.4 % crystal violet (CV) solution to the adhered cells in the wells and then stood at room temperature for 15 min. Excess stain was removed by rinsing the wells gently with 200 μL per using distilled water. This was repeated thrice. The microtiter plate was then air-dried for 1 h after, followed by addition of 200 μL of absolute ethanol to each well to solubilize the dye. The OD was measured at OD_590nm_ using a Safire Tecan-F129013 Microplate Reader (Tecan, Crailsheim, Germany). For each experiment, background staining was corrected by subtracting the crystal violet bound to un-treated controls (Blank) from those of the tested sample. The experiments were done in triplicate and average OD_590nm_ values were calculated. To estimate the antibiofilm activity (Abf A) of a given antibiotic the following equation was used, Abf A (%) = (1-(OD_Test sample_ - OD_Blank_)/ (OD _Untreated sample_ - OD _Blank_) × 100.

### Monitoring presence of relevant virulence factors of *P. aeruginosa spp*

#### (i) Detection of haemolysin

Haemolysin production by the *P. aeruginosa spp*. isolates were detected following protocols by Benson et al. [28]. The β-haemolytic activity was tested for on base agar (Himedia, India) supplemented with 7 % sheep erythrocytes for 18–24 h. Pure isolates were cultured on TSA, before streaking on blood agar and further incubated for 24 h at 37 °C. Zones of haemolysis around the colonies indicated the ability of these bacteria to haemolyse RBCs [40].

#### (ii) Detection of protease

To detect protease production by the *P. aeruginosa spp*. isolate skim milk agar was used and the protocol that was described in [28]. Briefly, two solutions (A and B) were made and used in this study. Solution A was prepared by adding 10 g skim milk to 90 ml of distilled water then volume was completed to 100 ml gently heated at 50 °C, then autoclaved and cooled to 50–55 °C. And solution B was also prepared by adding 2 g of agar powder to 100 mL of distilled water, mixed thoroughly, then autoclaved and cooled to 50–55 °C. Aseptically, 100 ml of solution A was mixed with 100 mL of solution B. Then the mixture was poured into sterile petri dishes, and then stored at 4 °C until use. This media used to detect the ability of the bacteria to produce protease [46]. The appearance of a cleared hydrolysis zone indicates a positive test [29].

#### (iii) Detection of lipase

Lipase production ability by *P. aeruginosa spp*. isolates were determined by methods outlined by Elliot et al. [29]. Briefly, a single colony of an overnight growth was cultured on Rhan medium, and then incubated for 1–5 days at 37 °C. The appearance of a turbid zone around colonies indicates a positive result [28].

#### (iv) Detection of lecithinase (phospholipase)

To detect lecithinase, we followed a standard procedure [30]. One pure colony was cultured on a medium of phospholipase activity assay followed by incubation for 1–3 days at 37 °C using established procedures [31]. The appearance of a white to brown colour elongated precipitated zone around colonies is considered a positive result [32].

### Validity and reliability

All experiments were conducted in triplicates that were independent of each other to validate reproducibility.

### Data analysis

Statistical analysis was performed using Graph pad Data on socio-demographics were summarized by frequencies and percentages. All values of diameter zones of inhibition are reported as mean ± standard error.

### Ethical consideration

Confidentiality and privacy were strictly adhered to and no names of individuals were recorded or made known in the reporting of information. The study was granted ethical clearance by the Institutional Research Ethics Committee (IREC) at Jaramogi Oginga Odinga Teaching and Referral Hospital (JOOTRH), written informed consent was obtained from the parent/guardian of each participate in this study.

## Results

Of 135 samples processed, 73 (54.1%) showed their growth as shown in **Supplementary data 1. A-F**. Among growth positive cases, 37(27.4%) were Gram positive while 36 (26.7%) were Gram negative and 62 (45.9%) did not show growths. High rate of growth positive rate in infection was found in case of male gender with Gram-positive cocci isolates at 23 (29.1%) and Gram-negative cocci isolates at 26 (32.9%) than female gender with Gram-positive isolates of 14 (25.5%) and Gram-negative isolates of 10 (18.2%), which were found to be statistically significant (p-value < 0.075). Among total growth, the highest growth rate was found in age groups >45 years in both Gram-positive and Gram-negative at 14 (26.9%) and 12 (23.1%) respectively. Least growth was found in an age group of < 5 years and 25-34 years as shown in **Table 1**.

**Table 1:** Sociodemographic characteristics of patients involved in the study for the period of May to August 2022

| Characteristic | Total (N, %) | Gram Positive (n, %) | Gram Negative (n, %) | No Growth | P Value |
| --- | --- | --- | --- | --- | --- |
| **Age Category** |  |  |  |  |  |
| < 5 years | 12 (9%) | 1 (8.3%) | 6 (50%) | 5 (41.7%) | 0.199 |
| 6-14 years | 24 (17.9%) | 9 (37.5%) | 6 (25%) | 9 (37.5%) |  |
| 15-24 years | 15 (11.2%) | 8 (53.3%) | 2 (13.3%) | 5 (33.3%) |  |
| 25-34 years | 16 (11.9%) | 2 (12.5%) | 6 (37.5%) | 8 (50%) |  |
| 35-44 years | 15 (11.2%) | 3 (20%) | 4 (26.7%) | 8 (53.3%) |  |
| >45 years | 52 (38.8%) | 14(26.9%) | 12 (23.1%) | 26 (50%) |  |
| Median Age (IQR) | 34 (IQR 13-56) | 25 (IQR 12-54) | 32.5 (IQR 12-53.5%) | 43 (IQR 22-59.5) |  |
| **Gender** |  |  |  |  |  |
| Female | 55 (41%) | 14 (25.5%) | 10 (18.2%) | 31 (56.4%) | 0.075 |
| Male | 79 (59%) | 23 (29.1%) | 26 (32.9%) | 30 (38%) |  |
| **Total** | **134** | **37 (27.6%)** | **36 (26.9%)** | **61 (45.5%)** |  |

Out of the total wound swab samples analyzed 71 (52.6%) were from swab while 64 (47.4%) were aspirate samples in which higher percentage growth was observed on aspirate than swab sample as in **Table 2**. Out of the 73(54.1%) growth positive wound swab samples,37 (27.4%) Gram-positive bacteria were found, while 36 (26.7%) Gram-negative bacteria were isolated. Among the Gram-positive isolates, *S. aureus* 28 (20.7%) was the most predominant followed by *Staphylococcus spp*. 9 (6.7%) while in Gram-negative isolates the most predominant organism was *Klebsiella spp*. 20 (14.8%) followed by *P. aeruginosa spp*. 10 (14.8%) and lastly *E. coli* at 6 (4.4%) as shown in **Table 2**.

**Table 2:** Distribution of pathogens by sample type and gram staining technique.

| Sample | Total (N, %) | *S. Aureus* | *Staphylococcus*<br>*spp.* | *Klebsiella*<br>*spp.* | *P.*<br>*Aeruginosa* | *E. coli* | No growth |
| --- | --- | --- | --- | --- | --- | --- | --- |
| Swab | 71(52.6%) | 9(12.7%) | 3(4.2%) | 3(4.2%) | 1(1.4%) | 0% | 55 (77.5%) |
| Aspirate | 64(47.4%) | 19(29.7%) | 6(9.4%) | 17(26.6%) | 9(14.1%) | 6(9.4%) | 7 (10.9%) |
| Total | 135(100%) | 28(20.7%) | 9(6.7%) | 20(14.8%) | 10(7.4%) | 6(4.4%) | 62 (45.9%) |

Among 37 (27.4%) Gram-positive isolates, 28 (20.7%) *S. aureus* resistance pattern showed that the most effective antibiotics were cotrimoxazole 13 (48.1%) followed by clindamycin 7 (25.9%) and erythromycin 7 (25.9%) while lower resistant to cotrimoxazole 3 (37.5%) was observed on *Staphylococcus spp*. as compared to *S. aureus*. In 36 (26.7%) Gram-negative isolates, 20 (14.8%) *Klebsiella spp*. resistance pattern showed that the most affected antibiotic was tetracycline 11 (61.1%) followed by gentamicin at 9(50%) while for *P. aeruginosa* four isolates show MRD to commonly used antibiotics, while for *E. coli* it was found that the isolates were sensitive to almost all of the common antibiotic used in the facility for their management as shown in **Table 3** and **Supplementary data 2. A-C**.

**Table 3:** Antibiotic resistant patterns of isolated pathogens

| Gram Positive | Cefoxitin | Oxacillin | Vancomycin | Penicillin | Cotrimoxazole | Clindamycin | Tetracycline | Gentamicin | Erythromycin |
| --- | --- | --- | --- | --- | --- | --- | --- | --- | --- |
| *S. aureus* | 3(11.1%) | 2(7.4%) | 2(7.4%) | 5(18.5%) | 13(48.1%) | 7(25.9%) | 5(18.5%) | 4(14.8%) | 7(25.9%) |
| *Staphylococcus*<br>*spp.* | 2(25%) | 0% | 1(12.5%) | 1(12.5%) | 3(37.5%) | 1(12.5%) | 1(12.5%) | 1(12.5%) | 1(12.5%) |
| Gram Negative | Cotrimoxazole | Ciprofloxacin | Cefuroxime | Ceftriaxone | Cefepime | Imipenem | Ampicillin | Gentamicin | Amikacin |
| *Klebsiella* spp. | 5 (27.8%) | 4 (22.2%) | 1 (5.6%) | 3 (16.7%) | 3 (16.7%) | 6 (33.3%) | 11(61.1%) | 9 (50%) | 2 (11.1%) |
| *P. aeruginosa* | 0% | 1(10%) | 2 (20%) | 1 (10%) | 1(10%) | 3 (33.3%) | 2 (20%) | 2 (20%) |  |
| *E. coli* | 0% | 0% | 0% | 0% | 0% | 1(16.7%) | 0% | 0% | 0% |

Among the total 10 (7.4%) *P. aeruginosa* isolates, 4 isolates (**12583**;**14421**;**1364 & 11985**) were found to have multi drug resistance. Among MDR isolates, 2 isolates (**12583 & 13642**) were resistant to 3 antimicrobial classes, 1 isolate (**13642**) was resistant to four antimicrobial classes; 2 isolates (**14421 & 13642** were resistant to five classes; 1 isolate (**11985**) was resistant to six antimicrobial Classes as shown in **Fig. 1**.

**Fig. 1:**
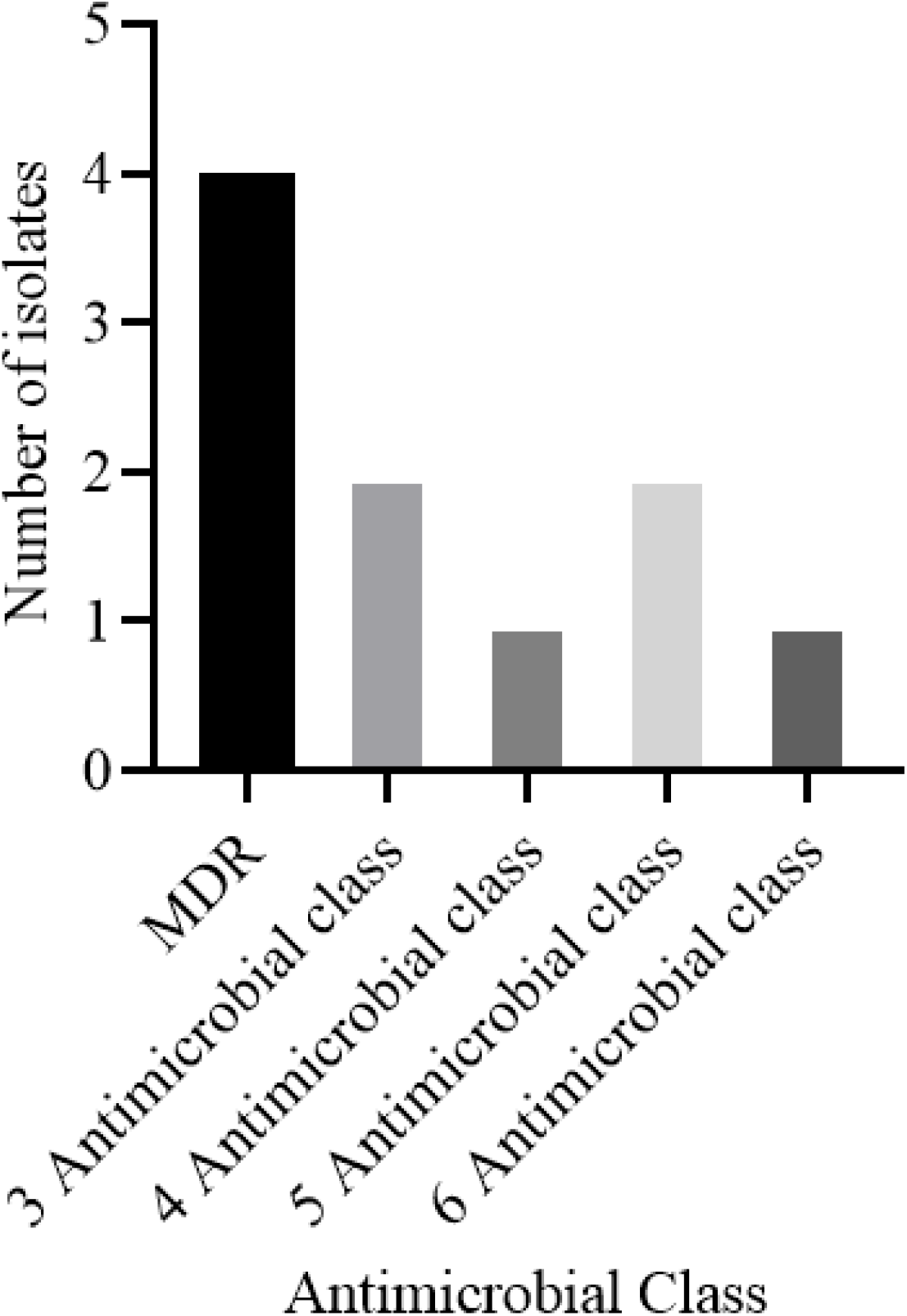
Multidrug resistance antimicrobial class resistant to P. *aeruginosa*.

This study investigated the production of various virulence enzymes like protease, phospholipase, lipase and haemolysin on the four *P. aeruginosa* isolates which showed MRD and were found to be resistant to common antibiotics used for its management within the study area. It was revealed that out of the 10 isolates four isolates showed resistance at different antimicrobial classes in which out of the four isolates, isolate **14421** was able to produce all types of the virulence enzymes. Also, it was confirmed that isolate **12583** was able to produce all enzymes except phospholipase enzyme while isolate **11985** was also capable of producing all except lipase enzyme. Lastly, it was determined that isolate **13642** was not able to produce all the virulence enzymes as shown in **Table 4**.

**Table 4:** Detection of some virulence factors of *P. aeruginosa*

| Isolates | Haemolysin | Protease | Lipase | Phospholipase |
| --- | --- | --- | --- | --- |
| *12583* | +p | +p | +p | -n |
| *13642* | -n | -n | -n | -n |
| *14421* | +p | +p | +p | +p |
| *11985* | +p | +p | -n | +p |

p = Positive and n = Negative

The isolates showed that the biofilm formation inhibitory effects of the various concentrations ml) −1(0.5, 0.25, 0.125, 0.0625 and 0.03125 mg were significantly lower than that of the positive control, an indication that biofilm formation was inhibited at these concentrations (**Figs. 2–4**). As much as such inhibitory effects were recorded, these findings clearly demonstrate that out of the four isolates that proved to be resistant to commonly used antibiotics, three isolates (12583; Fig.2,14421; Fig. 3 and 11985; Fig. 4) have the ability of forming biofilms.

**Fig. 2.**
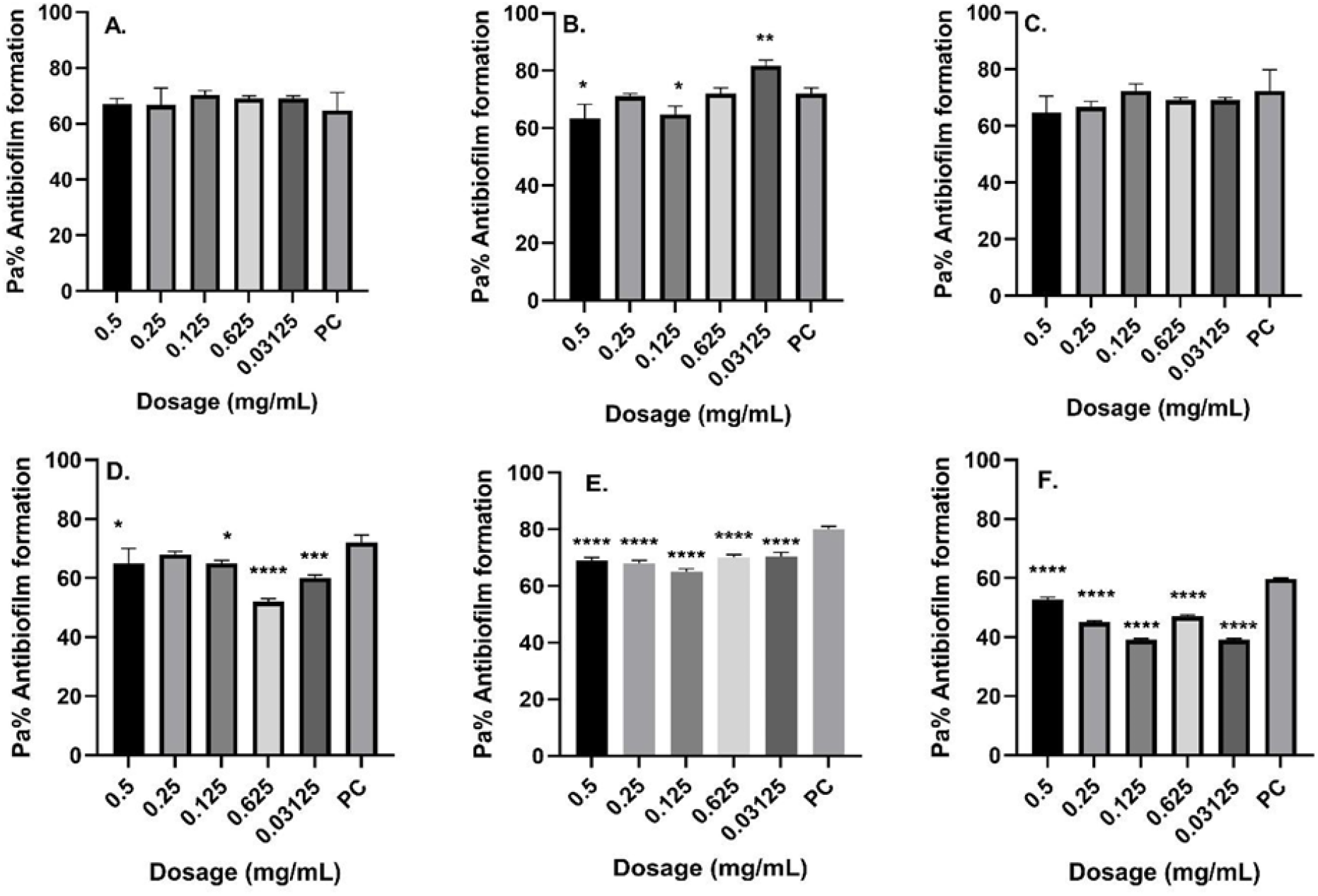
Antibiofilm formation activity against isolate **12583** of *P. aeruginosa* against various antibiotics: (a) Ciprofloxacin (b) Cefuroxime, (c) Ceftriaxone, (d) Ampicillin, (e) Gentamicin and (f) Amikacin; PC=*P. aeruginosa* ATCC^®^ 12934 *–* Positive control (*n*=3, ANOVA Dunnett’s multiple comparisons test; \**P*=0.05; \*\**P*=0.01; \*\*\**P*=0.001; \*\*\*\**P*=0.0001).

**Fig. 3.**
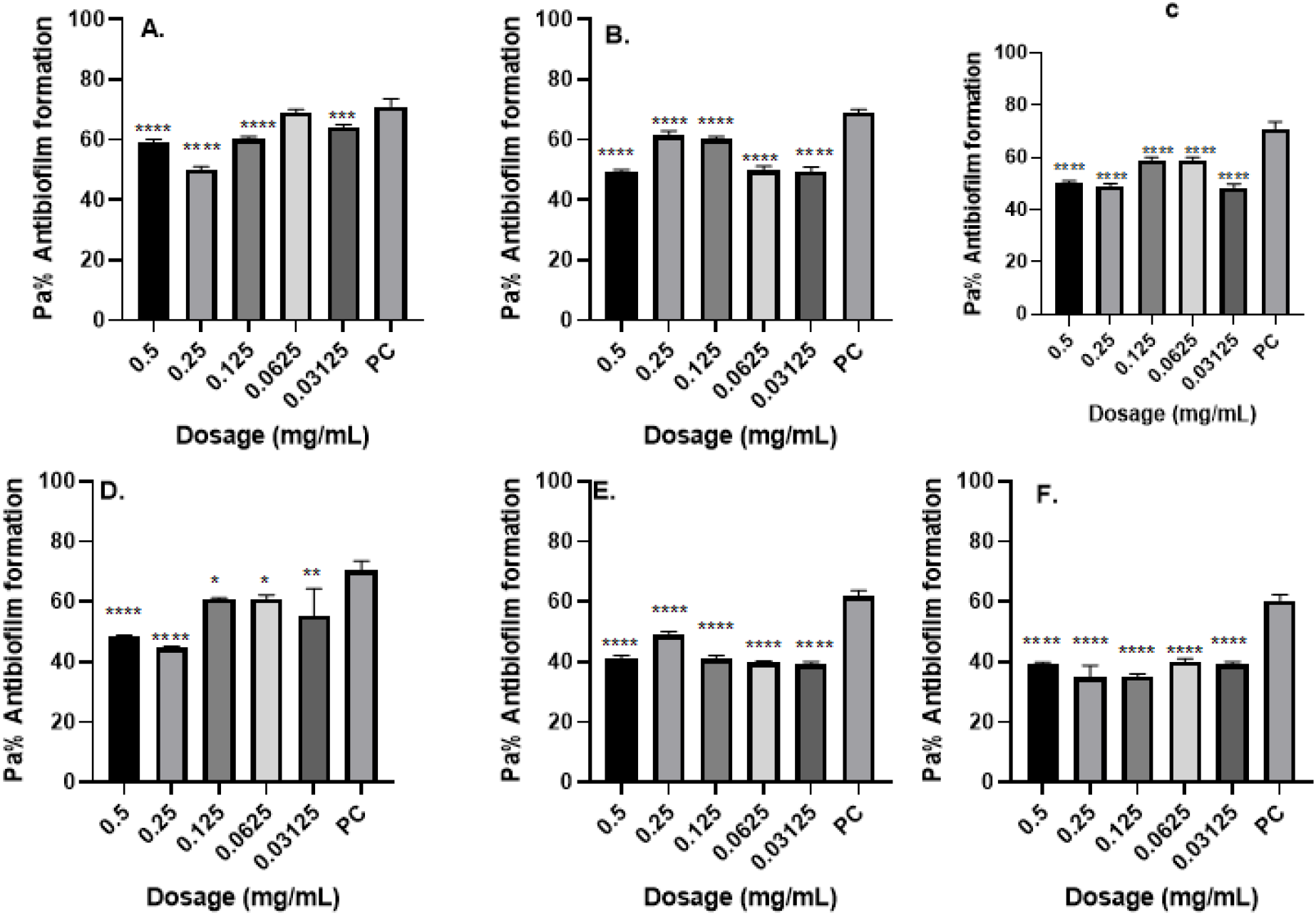
Antibiofilm formation activity against isolate **14421** of *P. aeruginosa* against various antibiotics: (a) Ciprofloxacin (b) Cefuroxime, (c) Ceftriaxone, (d) Ampicillin, (e) Gentamicin and (f) Amikacin; PC=*P. aeruginosa* ATCC^®^ 12934 *–* Positive control (*n*=3, ANOVA Dunnett’s multiple comparisons test; \**P*=0.05; \*\**P*=0.01; \*\*\**P*=0.001; \*\*\*\**P*=0.0001).

**Fig. 4.**
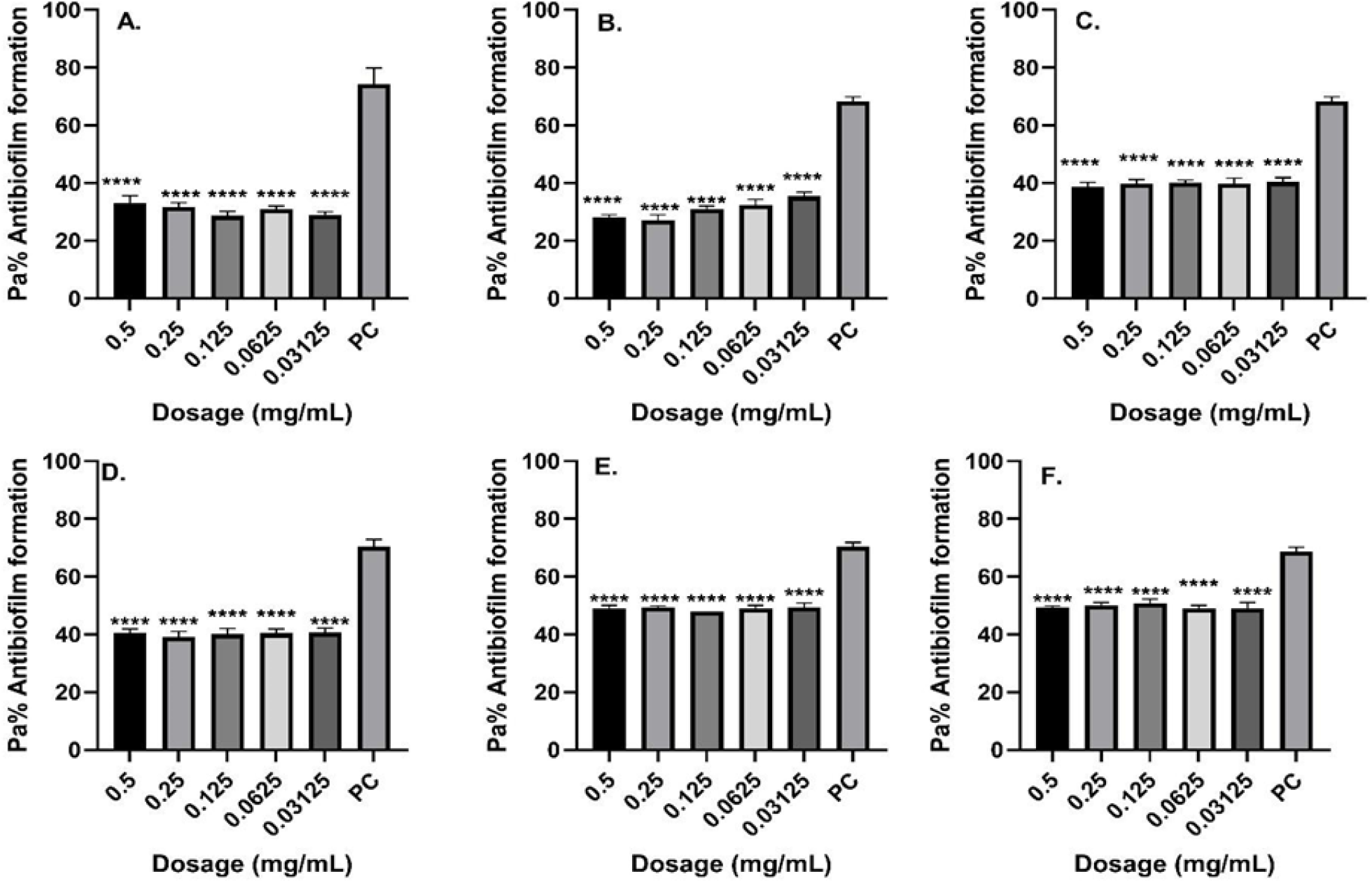
Antibiofilm formation activity against isolate **11985** of *P. aeruginosa* against various antibiotics: (A) Ciprofloxacin (B) Cefuroxime, (C) Ceftriaxone, (D) Ampicillin, (E) Gentamicin and (F) Amikacin; PC=*P. aeruginosa* ATCC^®^ 12934 *–* Positive control (*n*=3, ANOVA Dunnett’s multiple comparisons test; \**P*=0.05; \*\**P*=0.01; \*\*\**P*=0.001; \*\*\*\**P*=0.0001).

Antibiofilm formation activity against isolate 14421 of *P. aeruginosa* against various antibiotics like Ciprofloxine, Cefuroxime, Ceftriaxone, Ampicillin, Gentamicin and Amikacin against was observed in all the antibiotics (Fig. 4). Against ampicillin, a concentration of 0.0125 mg ml^−1^ and 0.0625 yielded significant biofilm formation inhibition (*P*=0.05), while for Gentamicin obtained significant differences on the biofilm formation inhibition at a concentration of 0.5 mg ml^−1^ (*P*=0.0001), 0.25 mg ml^−1^ (*P*=0.0001), 0.125 mg ml^−1^ (*P*=0.0001). 0.0625 mg ml^−1^ (*P*=0.0001) and 0.03125 mg ml^−1^ (*P*=0.0001) was observed in all the isolates. It is more worrying that Ciprofloxine, which is the commonly used antibiotic in the hospital, had less inhibitory effects against biofilm formation most so in the isolate 12583.

## Discussion

In this study, out of 135 samples from patients with wound infection, 73 (54.1%) samples showed positive growth. In similar studies conducted earlier by [22] in antibiotic susceptibility pattern of bacterial isolates causing wound infection among the patients visiting B & B Hospital in India, [25] showed growth positive was 44.9% & 50% respectively. A study conducted by [26,27] in the tertiary hospital in South Africa, more than 90% showed growth. Culture negative results observed in this study could be attributed to the non-infective bacteria, manual error in collection, transportation, overstaying of samples to reach Laboratory department culture media and collection of samples from patients taking antibiotics. The growth was seen to be higher in male patients in both Gram-positive and Gram-negative isolates at 23(29.1%) and 26(32.9%) respectively. A Similar study carried out by [24] agrees with our findings. The relatively higher percentage of male patients might be due to active involvement of males in outdoor activities, including agricultural work resulting in high infection possibility and prevalence of high rate of accidental cases among them [28].

From the total growth of 54.1%, the highest growth rate was found in age group >45 years followed by 6-14, 15-24, 25-34, 35-44 and lastly <5 years, our findings disagree with the studies carried out by [27,28] which showed highest growth rate in age group 21-30 (25.4%), followed by 31-40 (16.2%) and 11-20 (16.2%). Least growth was found in an age group of 0-10 and above 60 years. The reason behind this age group (>45) showing high growth among all age classes may be due to high prevalence of this age group in our study as well as the group being lively age group or working age group hence the chances of occurring accidents leading to hospital admission.

From the total bacterial isolates, 37(27.6%) were Gram positive and 36 (26.9%) were Gram negative bacteria. This result didn’t concur with a study conducted by [21-33] which shows Gram negative bacteria being predominant since their peptidoglycan layer is much thinner than that of Gram-positive bacilli and they are harder to kill because of their harder cell wall in which when their cell wall is disturbed release endotoxins that can make your symptoms worse [25]. Our observation of Gram negative as the most common bacteria in wound infections differs from other studies in Nigeria reporting Gram positive *to* be predominant [29]. Among the gram-negative isolated pathogens *Klebsiella spp*. was the most predominant 20(14.8%), The pathogenicity of *K. pneumoniae* is mediated by several virulence factors that allow it to evade host innate immune responses. These factors include the capsule, lipopolysaccharide, adhesins, iron acquisition systems, resistance to serum, and biofilm formation [24]. While for the Gram-positive S. *aureus* was the predominant isolate 28 (20.7%) and 1^st^ predominant isolates among the total. *S. aureus* pathogenicity is mediated by bacterial components and secreted virulence factors such as surface-associated adhesins, capsular polysaccharide (CP), and exotoxins [29]. This finding didn’t concur in a similar study conducted by [32], *P. aeruginosa* was the most predominant one (29.9%) among the total isolates this may be due to the permeability barrier afforded by its Gram-negative outer membrane, while *S. aureus* was predominant (27.5%) among Gram positive isolates. The study conducted by [32] Gram-negative bacilli constituted 66% of all the pathogens with *P. aeruginosa* as the most frequent (19%). Higher prevalence of *S. aureus* among Gram-positive bacteria and 1^st^ predominant isolates among the total seen in this study was also reported by other researchers, [35, 36]. Many researchers said that it is most commonly isolated. *S. aureus* was ubiquitous and the most common cause of localized suppurative lesions in human beings.

In the susceptibility patterns towards Gram-positive isolates, *S. aureus* isolates showed an alarmingly resistance to cotrimoxazole (48.1%), a commonly used antimicrobial drug in wound management Studies have shown *S. aureus* can become drug-resistant either via genetic mutations on DNA gyrase or through reduced expression of outer membrane proteins reducing drug accumulation [53]. Our findings corroborate previous studies [37] [38,39] that reported 47% and 49% resistance towards cotrimoxazole respectively. With cotrimoxazole has been the first-line treatment for wound infections, and it is commonly prescribed not only for wound infection but also for gastrointestinal tract infections, including diarrhea, but also for the treatment of respiratory tract infections, urinary tract infections and skin infections. Its mode of action is by targeting a subunit of DNA gyrase, which is essential in the production of bacteria DNA (39). It is often prescribed to immunocompromised patients, including human immunodeficiency virus (HIV) patients, which are common in the study area with a prevalence of 16.3% [38]. Such a vast usage of cotrimoxazole, and the fact that it is relatively cheap and could be acquired over the counter even without a prescription, may have contributed to the emergence of resistance towards this antibiotic as observed in this study.

A high incidence of Erythromycin resistant to *S. aureus* isolates (25.9%) was also observed. Erythromycin is a bacteriostatic and broad-spectrum antibiotic effective against a wide variety of Gram-positive bacteria. Its mode of action is by irreversibly binding to a receptor site on the 50S subunit of bacterial ribosome, inhibiting peptidyl transferase, consequently resulting in the prevention of amino acid transfer to growing peptide chains, leading to inhibition of protein synthesis. Up to late 2018, it was used as the first-line antibiotic treatment for typhoid and other *Salmonella* infections [32]. However, because of resistance and safety issues, it is no longer the first-line treatment in enteritis. In low-income countries, it is still widely used, as it is not expensive and readily available [40]. Erythromycin has been recommended by the World Health Organization (WHO) for the treatment of wound infection in both children and pregnant women [44]. The high resistance observed may be explained by its frequent usage for the treatment of severe cough and other infectious diseases. Also, Erythromycin resistance was found to be due predominantly to the presence of an Erm(B) methylase. Other reports have indicated that *S. aureus* isolates are resistant to Erythromycin [37]. A similar incidence was observed for other pathogens causing diarrhea in a study conducted in the study area [42].

A higher resistance rate of *Klebsiella spp*. to ampicillin (61.1%) was observed compared to that towards Gentamicin (50%), whose mode of activity is by targeting penicillin-binding proteins – a group of enzymes found anchored in the cell membrane, involved in the cross-linking of bacterial cell wall. *Klebsiella spp*. isolates from previous wound infections were known to exhibit resistance to ampicillin, [43] Gentamicin and Imipenem [43]. There was an emergence of *P. aeruginosa* isolates resistant to ampicillin (33.3%), Gentamicin, Amikacin and Ceftriaxone all at (20%), with 10 isolates resistant to this antimicrobial which has been primarily associated with the presence of IncC conjugative plasmids [44], hence leading to 4 isolates being found to be multi drug resistance. Among the MDR isolates, 2 showed resistance to 3 antimicrobial classes, 1 resistant to four antimicrobial classes; 2 resistant to five classes and finally 1 was resistant to six antimicrobial Classes. Studies on MRSA have shown their wide variation. Naik and Deshpande (2011) [45] showed 8.0% of MRSA which agree with our study. All *E. coli* isolates were sensitive towards all the antibiotics used except lower resistance to Imipenem, thus confirming the higher efficiency of these agents against *E. coli* isolates at Kisumu County.

Surprisingly, out of the four isolates, 3 (75%) isolates (**12583, 14421 & 11985**) were able to produce the haemolysin, a finding that didn’t conform with a previous study [46] which shows 100% isolates producing the enzyme. As stated before, purified haemolysin can cause fluid accumulation [32], in contrast to the watery fluid produced in response to CT, the accumulated fluid produced in response to haemolysin is invariably bloody with mucus [32]. Also, out of the four isolates 2 (50%) (**12583** and **14421**) had the ability to produce lipase. Our findings also concur with other studies, which showed that 50% isolates obtained in the study had the ability to produce lipase [16]. Lipase enzymes catalyze the hydrolysis of the ester bonds of triacylglycerols and may have a critical role in *P. aeruginosa* pathogenicity or nutrition acquisition. The production of an excess lipase allows bacteria to penetrate fatty tissue with the consequent formation of abscesses [48]. The production of these enzymes by the isolates may reflect the presence of genetic organization of a discrete genetic element, which encodes three genes responsible to produce proteases, lipases and phospholipase. This organization could be a possible part of a pathogenic island, encoding a product capable of damaging host cells and being involved in nutrient acquisition [49].

Some studies have suggested that Cefuroxime and Amikacin do inhibit growth at high concentrations [50, 51] but from this study, it was deduced that they inhibit biofilm formation at both lower and higher concentrations. The same scenario was also observed in most antibiotics like Ciprofloxine, Ampicillin, ceftriaxone and Gentamicin with such ‘Goldilocks’ effect. A possible explanation to the less activity observed at greater doses could be associated with the aggregation effects of the antibiotics at site of entry into bacteria cells especially at high dosages something that is not observed at lower dosages. It is likely that aggregation may favor biofilm formation as antibiotics struggle to reach the point of action and hence bacteria will continue to thrive and hence form more biofilms [52].

## CONCLUSION

The world is in dire need of improved antimicrobial owing to escalating AMR by a wide array of microbes on current antimicrobial pharmaceutics. This calls for a continuum of research efforts and innovation strategies aimed to catch up with AMR pace. This includes gene-mining applications, high-throughput genetic sequencing and molecular biology approaches, as well as, the recent incorporation of in silico tools and artificial intelligence (AI) in the antimicrobial search. That said, the collection of clinical AMR data remains pivotal in monitoring trends in antimicrobial resistance, in development of improved AMR drugs and in incorporation of innovative strategies such as AI. More focus is needed on the increasing bacterial antibiotic resistance especially in chronic wound management.

From this study, it can be concluded that antibiotic resistance patterns in Gram-positive and Gram-negative bacteria are in an alarming trend that leads to the failure of treatment. The resistance isolates do produce various virulence enzymatic factors such as haemolysin, lipase, protease and phospholipase, that are pivotal to aiding bacterial resistance. As much as inhibitory effects were recorded, these findings clearly demonstrate that the isolates proved to be resistant to commonly used antibiotics and they do form biofilms. And from the higher positive growth on the aspirant samples than the swab one we can conclude that the aspirant sample is the ideal sample for wound culture.

## Data Availability

All the data are shared in all the attached documents

## Acknowledgements

We thank JOOTRH Laboratories-Kisumu, Kenya for providing laboratory space and other resources used this study lastly, we would like to thank all staff in the microbiology section for the analysis of the samples.

## Author contributions

All authors contributed equally to this work.

